# Different predictors of message fatigue across diverse risk areas for dengue prevention in Taiwan

**DOI:** 10.1101/2024.11.26.24317959

**Authors:** Chia-Hsien Lin, Yen-Jung Chang, Hung-Yi Lu

**Affiliations:** Department of Health Promotion and Health Education, National Taiwan Normal University, Taipei City, Taiwan; Department of Communication & Graduate Institute of Telecommunications, National Chung-Cheng University, Chiayi County, Taiwan; School of Medicine, College of Medicine, National Sun Yat-Sen University, Kaohsiung City, Taiwan

## Abstract

**Background:** Different regions in Taiwan have varying levels of dengue risk; however, the same prevention messages were implemented nationwide for a prolonged period. People in areas with different risk levels may perceive the disease risk differently, and these perceptions may be linked to message fatigue. Individuals experiencing high levels of message fatigue may engage in fewer preventive activities. Although message fatigue has been studied concerning many infectious diseases, it has not been extensively researched in the context of dengue. Moreover, previous studies primarily focused on the general population regardless of risk level or on high-risk populations, with few examining message fatigue across areas with varying risk levels. Therefore, this study aimed to identify distinct predictors of message fatigue across different dengue-risk areas in Taiwan.

**Methodology/Principal Findings:** The online questionnaire was adapted from the measurement developed by So et al., based on the conceptual definition of message fatigue. Message fatigue has two dimensions: message environment (ME) and audience response (AR). A negative binomial model was used to examine the association between the demographic variables (sex, residency, education, occupation, age, and income) and the perceived variables (perceived prevalence, perceived severity, and optimistic bias) with ME and AR separately.

The results showed that in high dengue-risk areas, individuals with an education level of high school or below reported significantly higher levels of AR compared to those educated higher than high school (1.10, 95% CI = 1.01–1.21). In medium dengue-risk areas, individuals in the optimism group were 1.12 times more likely (95% CI = 1.01– 1.25) to report higher levels of ME than those in the realistic group. In low dengue-risk areas, younger individuals, males, and those with a high-school education or below reported significantly higher levels of AR.

**Conclusions/Significance:** This study revealed that dengue-message fatigue is associated with different variables across varying risk areas. These findings could inform global health communication strategies by identifying key demographic and perception-based predictors of message fatigue, enabling more targeted and effective public health messaging in regions with differing risk levels.

**Author summary:** In Taiwan, dengue is a significant public health concern, and different regions face varying levels of risk. Despite these differences, the same public health messages about dengue prevention are disseminated nationwide. In this study, we explored how people in areas with high, medium, and low risk of dengue of Taiwan respond to these messages, specifically focusing on message fatigue—when individuals tire of repeated health messages and may stop following preventive advice. We found that individuals in high-risk areas with lower education levels reported more fatigue with these messages, while those in medium-risk areas who were optimistic about the disease were more likely to experience fatigue. In low-risk areas, younger individuals, males, and those with lower education levels were more likely to experience message fatigue. These findings can help global health authorities design better-targeted public health messages by considering how different groups respond to repeated health messaging, thereby making dengue prevention efforts more effective across various risk areas.

## Introduction

Dengue is a significant communicable disease in Taiwan, with all suspected and confirmed cases reported to the Taiwan Centers for Disease Control (TW CDC) [1]. The entirety of Taiwan is susceptible to dengue due to two major mosquito vectors, *Aedes aegypti* and *Aedes albopictus* [1]. However, dengue incidence and prevalence vary across regions. The southern region, specifically Tainan, Kaohsiung, and Pingtung, accounted for over 98.6% of indigenous dengue cases between 1998 and 2023, followed by the northern, central, and eastern regions [1]. Studies have shown the highest prevalence rates were also in the southern region, while the central region generally had the lowest rates [2] [3]. Therefore, in this study, the southern, northern, and central regions served as proxies for high, medium, and low dengue risks, respectively. Throughout the text, we will refer to these regions as high, medium, and low-risk areas.

The current prevention and control strategies were established in 2003 [1]. These are maintained throughout the year and consist of information, education, and communication (IEC) activities, as well as routine vector control. The TW CDC conducts IEC activities, which include health education campaigns through television advertisements, radio announcements, posters, the Internet, and social media platforms.

These activities are uniform across Taiwan. They aim to increase public awareness of self-protection measures, symptom recognition, and the management of water-holding containers on private properties. However, dengue continues to occur each year despite year-round IEC activities, suggesting that these efforts may need modification to find new approaches to motivate people to undertake prevention measures.

One reason that may reduce motivation to participate in disease prevention is message fatigue [4-6]. Message fatigue is a psychological state from prolonged exposure to similar or repetitive messages, resulting in negative reactions such as boredom, annoyance, and disengagement [7]. When individuals are overwhelmed with too many messages on the same topic, they may become mentally exhausted and bored, leading to message fatigue [8]. This state encompasses perceived overexposure, redundancy, exhaustion, and tedium [4, 7, 8]. Based on So et al.’s study, message fatigue consists of the dimensions of message environment (ME) and audience response (AR). The ME dimension describes a sense of being overwhelmed and frustrated by excessive and repetitive information about the message. The AR dimension indicates boredom and annoyance with continuous and uninteresting messages [7].

Several demographic factors, such as age, sex, education, job, and income level, have been identified as being associated with message fatigue [9-12]. However, the patterns of these demographic factors are not consistent across studies. For example, Stockman et al. aimed to examine the level of fatigue toward HIV-prevention messages among three high-risk populations: men who have sex with men, heterosexuals, and injection drug users (IDUs). The findings showed that younger IDUs had a higher level of HIV-prevention fatigue than older IDUs did [9]. In contrast, Steiner et al. aimed to determine whether the volume of automated text or interactive voice response messages influenced the likelihood of patients opting out of future messages, finding that the senior group (aged 55 and older) experienced a higher level of message fatigue than younger individuals (aged 18–34) did [10]. Regarding sex, a Polish study found that women reported significantly greater COVID-19 fatigue, particularly in terms of information fatigue [12], while men had higher levels of anti-tobacco message fatigue in the United States [11].

Message fatigue has been observed in prevention messages for infectious diseases [5, 8, 9, 13-15]. For example, research by Guan et al. examined the impact of COVID-19 message fatigue on people’s intentions to engage in preventive behaviors [5]. The results showed that message fatigue leads to both reactance and inattention, which in turn negatively affect people’s intentions to engage in preventive behaviors, mediating the relationship between message fatigue and these behavioral intentions [5]. Regarding HIV prevention messages, Stockman et al. aimed to examine HIV-prevention fatigue in community-based samples of three high-risk populations in San Francisco. The study found that IDU participants had a higher level of fatigue toward HIV-prevention messages compared to heterosexuals [9]. A report by UNICEF’s Eastern and Southern Africa Regional Office showed that a prolonged period of community awareness regarding Ebola prevention resulted in message fatigue [15]. However, few studies on message fatigue have been conducted in the context of dengue.

Moreover, most studies on prevention messages for infectious diseases focus on the general population regardless of their risk levels or solely on high-risk populations concerning message fatigue. Few studies have evaluated message fatigue across groups with different risk levels, such as individuals residing in various areas but exposed to the same prolonged messaging. Groups with different risk levels may perceive disease risk differently, influenced by factors such as psychological and socio-economic conditions [16-18]. Additionally, perceptions of disease risks are associated with message fatigue [7, 8]. Therefore, groups with different risk levels may experience varying levels of fatigue.

To inform future message designs, this study aimed to identify distinct predictors of message fatigue across various dengue-risk areas in Taiwan. The examined predictors included demographic variables such as age, sex, education, and income, as well as perceived risk variables, including optimistic bias, perceived prevalence, and perceived severity. This study could inform global health communication strategies by identifying key demographic and perception-based predictors of message fatigue, allowing for more targeted and effective public health messaging in diverse regions at varying risk levels.

## Materials and methods

### Participants and procedure

The study was part of a larger project involving an online survey on dengue perceptions and dengue-message fatigue. Participation was voluntary, and respondents were assured of confidentiality and anonymity. Participants were adults aged 18 and above residing in Taiwan, who reported their demographic information, perceived risks, and responses to questions assessing dengue-message fatigue. The questionnaire was distributed through the authors’ social media networks, including Facebook and Line. A convenience sampling method was employed. The survey started on October 25, 2023, and ended on November 13, 2023.

### Measurements of variables

#### Message fatigue

The online questionnaire was adjusted based on the measurement developed by So et al. according to the conceptual definition of message fatigue [7]. Similar to the study by So et al., message-fatigue measurement included the ME and AR dimensions. Unlike the original study, in this study, the ME dimension consisted of six questions to represent overexposure and redundancy (S1 Table), with a Cronbach’s alpha of 0.85. The reduction from the original nine questions to six was due to the translation process from English to Chinese, during which it was found that some questions could be combined or simplified without losing their original meaning. Therefore, the number of questions was reduced while maintaining the consistency and clarity of the content. For the AR dimension, the number of questions was reduced from seven to five, representing exhaustion and tedium in this study, for the same reason (S1 Table), with a Cronbach’s alpha of 0.88. All questions were measured on a 5-point Likert scale ranging from 1 (strongly disagree) to 5 (strongly agree) [7].

#### Perceived risks

Perceived risks included perceived prevalence, perceived severity, and optimistic bias. Perceived prevalence was assessed with the question, “Do you think the current dengue outbreak in Taiwan is severe?” and perceived severity with, “Do you think the likelihood of death is high once infected with dengue?” Both were measured on 11-point Likert scales ranging from 0 (strongly disagree) to 10 (strongly agree).

Optimistic bias was evaluated with two questions: “How likely do you think it is that you will have a dengue infection?” and “How likely do you think it is that others around you will have a dengue infection?” These two questions were measured on 11-point Likert scales ranging from 0 (strongly unlikely) to 10 (strongly likely). The score of the first question was subtracted from that of the second to categorize optimistic bias into three groups: realistic (score zero), optimistic (positive score), and pessimistic (negative score).

#### Demographic variables

Sex, residency, education, job, age, and income were the demographic variables. Sex was categorized as male and female. Residency was categorized into five regions: north, central, south, east, and outlying islands. Education was classified into two levels: higher than high school and equal to or below high school. Job experience was categorized as no job experience in dengue control and relevant job experience. Age and average household monthly income were continuous variables.

#### Statistical analysis

The dimensions of ME and AR were assessed separately, as the questions described different feelings. The independent variables were demographic factors and perceived risks. To identify the most suitable model, supervised backward selection with a χ^2^ test at a 0.05 significance level was used with Poisson and negative binomial models. The Akaike Information Criterion (AIC) determined the final model, based on the lowest value. All model selections and tests were in R version 4.2.2 [19, 20].

#### Ethics Statement

This study utilized an online questionnaire that was previously reviewed and approved by the Research Ethics Committee (REC) of National Taiwan Normal University under approval number 202312HS019. As this study was conducted anonymously, no personal identifying information was collected through the online platform, ensuring participants’ privacy and confidentiality. Participants were informed about the purpose of the research at the beginning of the online questionnaire, and their voluntary participation was implied by their completion of the survey. The study adhered to ethical principles for research involving human participants, particularly with respect to anonymity and the protection of data collected through online methods.

## Results

During the survey, 818 individuals completed it and responded validly. Participants under the age of 18 (N = 4) and those residing in the eastern (N = 10) and outlying islands (N = 3) were excluded. The exclusions based on residency in the eastern and outlying islands were for statistical considerations. Consequently, the final sample consisted of 801 individuals.

Among the participants, females (N = 531) were more than males (N = 270). Most respondents had a higher education level than high school (N = 677), while a smaller portion had an education level equal to or below high school (N = 124). Additionally, a larger group reported no job experience in dengue control (N = 683) compared to those with relevant job experience (N = 118). In terms of residency, participants were distributed across the south (N = 466), central (N = 116), and north (N = 219) regions. The age range of participants was from 18 to 88 years, with an average age of 46.3 years. Both perceived prevalence and perceived severity of dengue ranged from 0 to 10, with mean scores of 6.5 and 5.0, respectively. Regarding the optimism bias variable, the majority of respondents were classified as realistic (N = 533), followed by those in the optimism (N = 205), and pessimism groups (N = 63).

### High dengue risk (i.e., southern region)

The total number of participants in the high dengue-risk region was 466. More participants were female (N = 310), had a higher education level than high school (N = 365), and lacked job experience in dengue control (N = 384), when compared to males (N = 156), those with an education level equal to or below high school (N = 101), and those with job experience in dengue control (N = 82). Participants’ ages ranged from 18 to 83 years, with an average age of 49.4 years. In terms of perceived risks, both perceived prevalence and perceived severity of dengue ranged from 0 to 10, with mean scores of 6.9 and 5.0, respectively. The optimism bias variable showed that the largest proportion of participants were in the realistic group (N = 308), followed by the optimism (N = 121), and pessimism groups (N = 37).

For the ME dimension in the southern region, due to a lower AIC, the negative binomial model (AIC = 3063.30) was used instead of the Poisson model (AIC = 3257.76) for evaluating the associations. By applying the negative binomial model, no specific significant predictors were identified for the feelings of boredom and annoyance associated with dengue messages in southern Taiwan (Table 1).

**Table 1.** Negative binomial model for dengue-message fatigue considering message environment and audience response in southern Taiwan (N = 466).

| Southern region | Predictors | Est | SE | Exp(Est) | 95% CI<sup>a</sup> |
| --- | --- | --- | --- | --- | --- |
| ME<sup>b</sup> | - | - | - | - | - |
| AR<sup>c</sup> | Education (equal to or below H vs. higher H) | 0.099 | 0.048 | 1.104 | 1.005 - 1.213 |

<sup>a</sup>95% CI referring to Exp(Est)
<sup>b</sup>Message environment
<sup>c</sup>Audience response

Given the lower AIC, the negative binomial model (AIC = 2891.50) was chosen over the Poisson model (AIC = 3112.21) to evaluate associations related to message fatigue in the AR of the southern region. The model indicated that individuals with an education level equal to or below high school reported significantly higher levels of boredom and annoyance compared to those with a higher education level (1.10, 95% CI = 1.01–1.21) in southern Taiwan (Table 1).

### Medium dengue risk (i.e., northern region)

The region of medium dengue risk had 219 participants. Among them, females (N = 148) outnumbered males (N = 71). A lesser percentage of respondents (N = 7) had education levels equal to or below high school, whereas the majority (N = 212) had higher education levels. Furthermore, compared to those with relevant job experience (N = 13), a larger group (N = 206) reported no prior experience in dengue control. Participants ranged in age from 18 to 88 years, with an average age of 39.5. With average scores of 5.8 and 4.8, respectively, the perceived severity and prevalence of dengue ranged from 0 to 10. A large proportion of respondents (N = 153) were categorized as realistic regarding the optimism bias variable, followed by those in the optimism (N = 52) and the pessimism groups (N = 14).

Due to a lower AIC for message fatigue when considering the ME, the negative binomial model (AIC = 1351.00) was applied rather than the Poisson model (AIC = 1392.81) to evaluate the association. The model showed that the optimistic bias variable (p = 0.027) was the only predictor. The result indicated that the optimism group was 1.122 times more likely (95% CI = 1.011–1.246) to feel frustrated by a large number of dengue messages than the realistic group. However, compared to the realistic group, the pessimism group did not feel significantly more frustrated with the overabundance of dengue messages (Table 2).

**Table 2.**
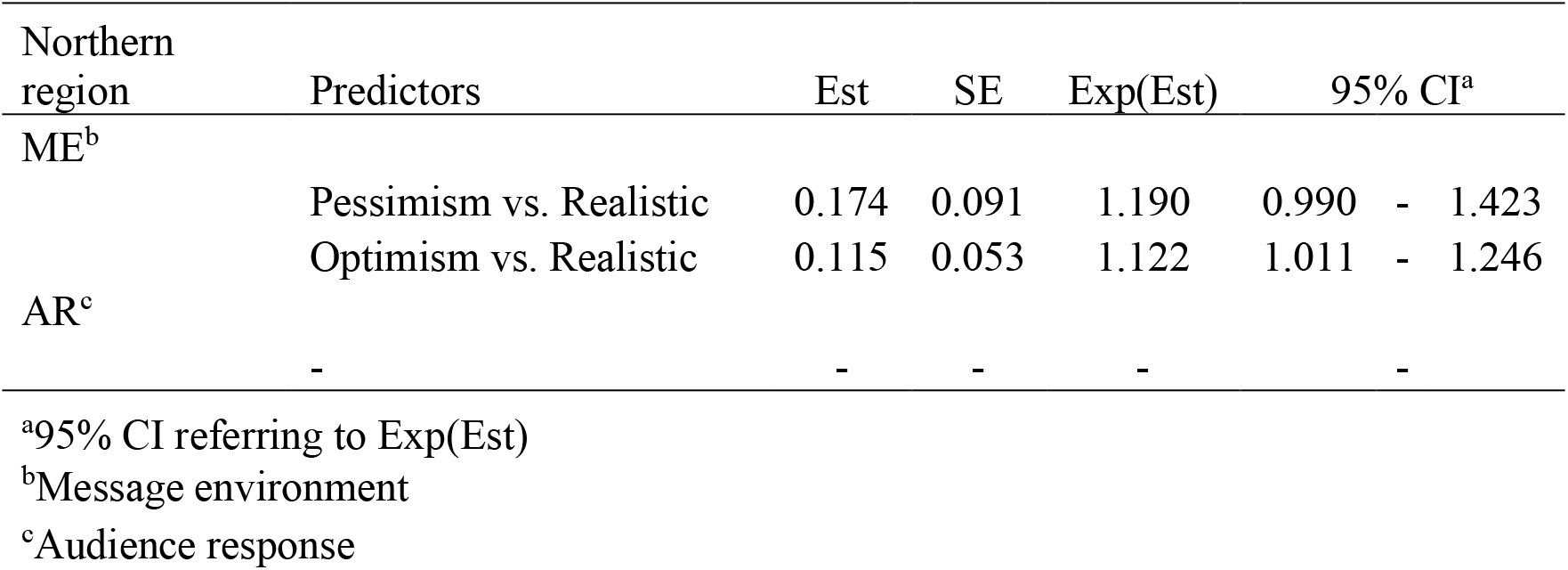
Negative binomial model for dengue-message fatigue considering message environment and audience response in northern Taiwan (N = 219).

Regarding AR, the negative binomial model (AIC = 1306.28) was chosen instead of the Poisson model (AIC = 1390.88). However, applying the negative binomial model did not identify any significant predictors for message fatigue in the AR of the north region (Table 2).

### Low dengue risk (i.e., central region)

In the region with low dengue risk, more participants were female (N = 73), had higher education levels than high school (N = 100), and lacked job experience in dengue control (N = 93), when compared to males (N = 43), those with education levels equal to or below high school (N = 16), and those with job experience in dengue control (N = 23). Ages ranged from 18 to 79 years, with an average of 45.8 years. Perceived risks of dengue showed both prevalence and severity ranging from 0 to 10, with mean scores of 6.4 and 5.0, respectively. The optimism bias variable indicated that the majority of participants fell into the realistic group (N = 72), followed by the optimism group (N = 32), and the pessimism group (N = 12).

For ME in the central region, due to a lower AIC, the negative binomial model was chosen (AIC = 735.75) instead of the Poisson model (AIC = 639.12) for evaluating the associations. Significant predictors were age (p = 0.027), sex (p = 0.033), and education (p = 0.010). Younger individuals (0.995, 95% CI = 0.991–0.999), males (1.145, 95% CI = 1.010–1.298), and people with education levels equal to or below high school (1.255, 95% CI = 1.058–1.489) felt more frustrated with an overabundance of dengue messages compared to older individuals, females, and people with higher education levels (Table 3).

**Table 3.**
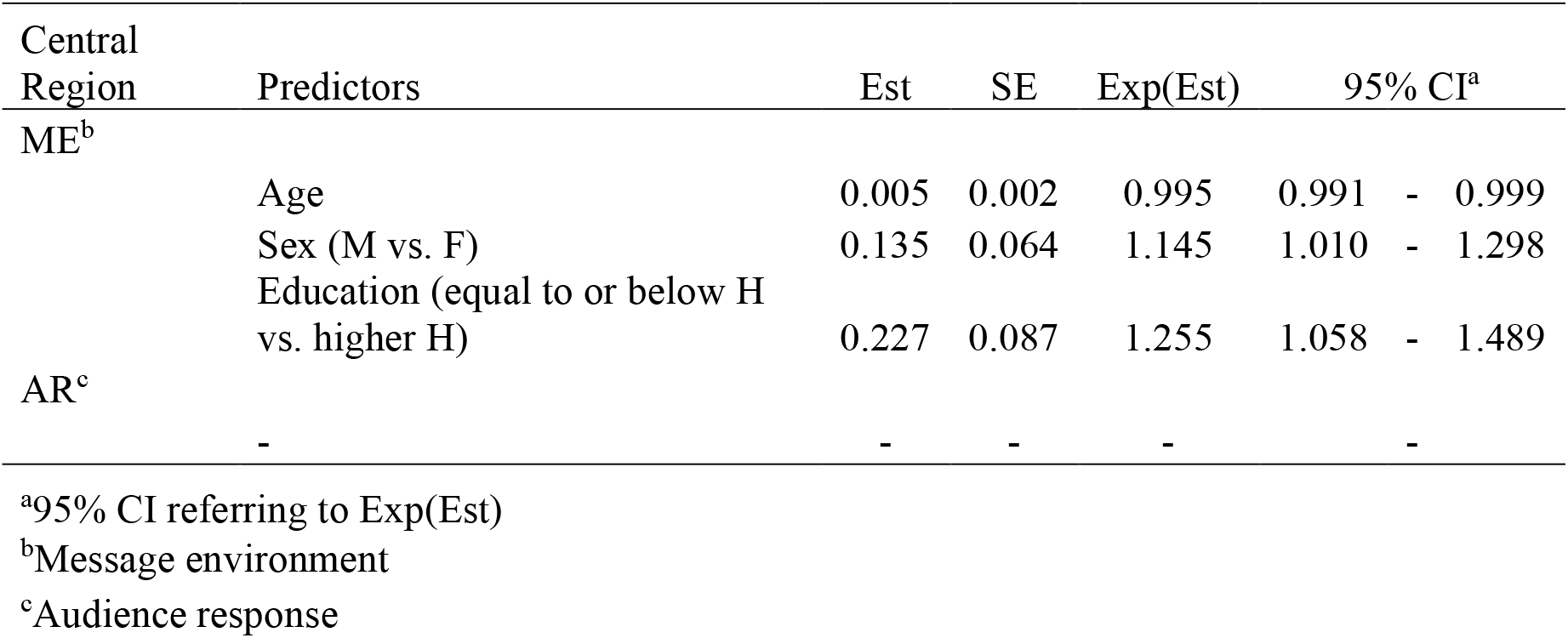
Negative binomial model for dengue-message fatigue considering message environment and audience response in central Taiwan (N = 116).

It was decided to use the negative binomial model (AIC = 706.53) rather than the Poisson model (AIC = 759.88) to assess associations related to message fatigue in the AR dimension due to the lower AIC. However, by applying the negative binomial model, no significant predictors for message fatigue in the AR of the central region were identified (Table 3).

## Discussion

In this cross-sectional study, we assessed the two dimensions of message fatigue across different regions, which represented varying levels of dengue risk, and analyzed them in relation to demographic factors and perceived risks. The study found that predictors of message fatigue varied across regions with different levels of dengue risk. In the high dengue-risk region, no significant predictors were identified for the ME dimension, while education was a significant predictor for AR. Combining ME and AR, in the high dengue-risk region, people with an education level equal to or below high school had higher message fatigue compared to those with a higher education level (above high school). In the medium dengue-risk region, optimism bias was the only predictor for the ME dimension, and AR had no predictor. This indicates that people in the optimism group were more likely to experience dengue-message fatigue compared to those in the realistic group. In the low dengue-risk region, age, sex, and education were significant predictors for ME, while no significant predictors were identified for the AR dimension. This means that younger individuals, males, and people with an education level equal to or below high school had a higher level of message fatigue.

This study showed that individuals with lower education levels had higher dengue-message fatigue in the southern and central regions, which represented high and low dengue risks, respectively. The tendency for individuals with lower education levels to experience higher health-message fatigue could be attributed to several factors. One factor is that individuals with lower education levels may have less prior knowledge or experience with health-related topics. According to Cognitive Load Theory [21], the cognitive effort required to process, understand, and integrate new health information is greater for these. This increased cognitive load may limit their ability to fully comprehend and engage with health messages, leading to mental exhaustion, confusion, and fatigue [21, 22]. Another factor could be psychological exhaustion. The psychological impact of prolonged exposure to health messages can cause burnout.

Individuals with low education levels might not have the coping mechanisms [23, 24] or support systems [23, 25] to process ongoing health communications effectively. Therefore, they may experience this fatigue more acutely. To mitigate their fatigue, simple [21], relevant [26], and engaging [27] dengue messages could be considered. Younger individuals in low dengue-risk central Taiwan exhibited a higher level of dengue-message fatigue in this study. While this phenomenon has not been specifically observed in dengue messaging, it has been seen in other types of messages, such as anti-tobacco [11] and COVID-19 vaccine booster uptake [28]. The tendency of younger individuals to experience greater message fatigue may be because they, especially those active on social media and other digital platforms [29], are often exposed to a higher volume of information than older individuals are. Repeated exposure to the same or similar messages, such as those related to dengue, can lead to saturation and fatigue when the messages no longer feel relevant or urgent. Additionally, younger people often perceive themselves as less vulnerable to diseases [30, 31]. If they believe they are not at significant risk, they may view repeated messages as unnecessary or irrelevant, leading to disengagement and fatigue. Furthermore, in Taiwan, the way dengue prevention messages are framed or delivered might not resonate well with younger audiences. If the communication does not align with their interests, values, or preferred communication channels, it can lead to disinterest and fatigue [32, 33]. Messages could be designed in formats and delivery channels that align with younger people’s digital habits and interests [34, 35]. By making the messages feel fresh, relevant, and interactive, the campaign can better combat message fatigue and effectively engage this group [36-39].

In addition to younger individuals, males were also identified as predictors of dengue-message fatigue in central Taiwan (i.e., low dengue risk), compared to females. This finding is interesting because most studies report higher message fatigue in females [12, 40, 41], with fewer examples of fatigue in males [11]. In this study, males exhibited a higher level of message fatigue, which could be due to the fact that males are less likely than females to seek out health information actively in Taiwan [42, 43]. Therefore, when males are exposed to health messages, they might find them more overwhelming, leading to quicker fatigue. Another possible explanation is that males may not engage with health messages on an emotional level as deeply as females do [44, 45]. This lack of engagement could lower the threshold for perceiving messages as repetitive or unimportant, thus increasing fatigue. Males may also be more resistant to certain types of health messaging, particularly those that challenge their independence or self-reliance. For example, messages that encourage using mosquito repellents, wearing long sleeves, or using bed nets might be seen as an imposition on their usual routines or as suggesting that they cannot protect themselves without these aids. This resistance can increase fatigue because they might perceive these messages as unnecessary.

Individuals in the optimism group were more prone to experiencing dengue-message fatigue compared to those in the realistic group in northern Taiwan (i.e., medium dengue risk), according to this study. In the optimism group, people tend to believe they are less likely to be affected by negative outcomes compared to others, such as getting infected with dengue [46, 47]. Therefore, even in areas with high prevalence, they may think, “It won’t happen to me,” lowering the perception of personal risk. In our sample, people living in the northern region had the lowest level of perceived prevalence for dengue compared to other regions, supporting this idea. This optimism bias results in message fatigue because it causes individuals to perceive health messages as irrelevant or unnecessary. This perception reduces their engagement with the messages and increases their resistance to the content, leading to fatigue. Another possibility is that optimistic individuals may believe that their actions are sufficient to protect them, leading them to disregard additional advice or warnings. This overconfidence reduces their receptivity to ongoing messages, causing fatigue [48]. For this group, messages framed around potential losses could be more impactful for those who might feel overly optimistic about their situation. In other words, if messages focus on what they could lose, such as their health, well-being, or the safety of their loved ones if they don’t act, it could help them take the situation more seriously and avoid being too relaxed about it.

This study demonstrated different demographic and perception-based predictors for message fatigue across regions with varying risk levels. This implies that messages must be tailored to specific risk regions to achieve effective health communication. However, this study has some limitations. First, while message fatigue was the focus of this study, which could reduce motivation to participate in disease prevention, another possibility that could also lower motivation is the boomerang effect. This effect occurs when people believe their freedom is at risk, leading them to resist the advice of a message. Second, this survey used convenience sampling. While convenience sampling is easy to use and inexpensive, making it beneficial in exploratory research, it is a nonprobability sampling technique. Therefore, the samples in this study might not accurately reflect the entire population (all Taiwanese citizens aged 18 years or older). Third, the survey was conducted using an online questionnaire. Respondents might misunderstand questions, leading to inaccurate or inconsistent data. To minimize this misunderstanding, various individuals pre-tested the questionnaire. Lastly, these findings can identify associations between message fatigue and other variables, but they cannot establish causality.

## Data Availability

All relevant data are within the manuscript and its Supporting Information files.

## Supporting information

**S1 Table. Dimensions and questions of message fatigue. Questions adapted from So et al**.

**S2 Table. Dataset**

**S3 Table. Variables, codes and categories in the dataset**

## References

1. Centers for Disease Control ROCT. Guidelines for Dengue / Chikungunya Control 2024 [Chinese]. Taiwan: Centers for Disease Control, R.O.C. (Taiwan); 2024.

2. Hsu JC, Hsieh CL, Lu CY. Trend and geographic analysis of the prevalence of dengue in Taiwan, 2010-2015. Int J Infect Dis. 2017;54:43–9. doi: doi.org/10.1016/j.ijid.2016.11.008.

3. Lee Y-H, Hsieh Y-C, Chen C-J, Lin T-Y, Huang Y-C. Retrospective Seroepidemiology study of dengue virus infection in Taiwan. BMC Infectious Diseases. 2021;21(1):96. doi: 10.1186/s12879-021-05809-1.

4. Mao B, Jia X, Huang Q. How do information overload and message fatigue reduce information processing in the era of COVID-19? An ability–motivation approach. J Inf Sci. 2022. doi: doi.org/10.1177/01655515221118047.

5. Guan M, Li Y, Scoles JD, Zhu Y. COVID-19 message fatigue: How does it predict preventive behavioral intentions and what types of information are people tired of hearing about? Health Communication. 2023;38(8):1631–40. doi: 10.1080/10410236.2021.2023385.

6. Jia X, Ahn S, Carcioppolo N. Measuring information overload and message fatigue toward COVID-19 prevention messages in USA and China. Health Promot Int. 2023;38(3). Epub 2022/01/30. doi: 10.1093/heapro/daac003.

7. So J, Kim S, Cohen H. Message fatigue: Conceptual definition, operationalization, and correlates. Communication Monographs. 2017;84(1):5–29. doi: 10.1080/03637751.2016.1250429.

8. Sun J, Lee SK. “No more COVID-19 messages via social media, please”: the mediating role of COVID-19 message fatigue between information overload, message avoidance, and behavioral intention. Current Psychology. 2023;42(24):20347–61. doi: 10.1007/s12144-023-04726-7.

9. Stockman JK, Schwarcz SK, Butler LM, de Jong B, Chen SY, Delgado V, et al. HIV prevention fatigue among high-risk populations in San Francisco. J Acquir Immune Defic Syndr. 2004;35(4):432–4. Epub 2004/04/21. doi: 10.1097/00126334-200404010-00016.

10. Steiner JF, Zeng C, Comer AC, Barrow JC, Langer JN, Steffen DA, et al. Factors associated with opting out of automated text and telephone messages among adult members of an integrated health care system. JAMA Network Open. 2021;4(3):e213479–e. doi: 10.1001/jamanetworkopen.2021.3479.

11. So J, Popova L. A profile of individuals with anti-tobacco message fatigue. American journal of health behavior. 2018;42(1):109–18.

12. Domosławska-Żylińska K, Krysińska-Pisarek M, Włodarczyk D. Gender-specificity of fatigue and concerns related to the COVID-19 pandemic-A report on the Polish population. Int J Environ Res Public Health. 2023;20(7). Epub 2023/04/14. doi: 10.3390/ijerph20075407.

13. Macapagal K, Birkett M, Janulis P, Garofalo R, Mustanski B. HIV prevention fatigue and HIV treatment optimism among young men who have sex with men. AIDS Educ Prev. 2017;29(4):289–301. Epub 2017/08/22. doi: 10.1521/aeap.2017.29.4.289.

14. Kim JH, Kwok KO, Huang Z, Poon PK, Hung KKC, Wong SYS, et al. A longitudinal study of COVID-19 preventive behavior fatigue in Hong Kong: a city with previous pandemic experience. BMC Public Health. 2023;23(1):618. Epub 2023/04/03. doi: 10.1186/s12889-023-15257-y.

15. Esaro U. Risk Communication and community engagement for Ebola virus disease preparedness and response-lessons learnt and recommendations from Burundi, Rwanda, South Sudan, Tanzania and Uganda 2020. Available from: https://www.unicef.org/esa/reports/risk-communication-and-community-engagement-ebola-virus-disease-preparedness-and-response.

16. Yin JD-C, Lui JN-M. Factors influencing risk perception during Public Health Emergencies of International Concern (PHEIC): a scoping review. BMC Public Health. 2024;24(1):1372. doi: 10.1186/s12889-024-18832-z.

17. Vornanen M, Konttinen H, Peltonen M, Haukkala A. Diabetes and cardiovascular disease risk perception and risk indicators: A 5-Year follow-up. International Journal of Behavioral Medicine. 2021;28(3):337–48. doi: 10.1007/s12529-020-09924-2.

18. Adachi M, Murakami M, Yoneoka D, Kawashima T, Hashizume M, Sakamoto H, et al. Factors associated with the risk perception of COVID-19 infection and severe illness: A cross-sectional study in Japan. SSM Popul Health. 2022;18:101105. Epub 2022/05/03. doi: 10.1016/j.ssmph.2022.101105.

19. Venables WN, Ripley BD. Modern Applied Statistics with S. Fourth ed: Springer; 2002.

20. Team RC. R: A language and environment for statistical computing. R Foundation for Statistical Computing, Vienna, Austria. URL https://www.R-project.org/ 2022.

21. Sweller J. Cognitive load theory, learning difficulty, and instructional design. Learning and Instruction. 1994;4(4):295–312. doi: doi.org/10.1016/0959-4752(94)90003-5.

22. Cutler DM, Lleras-Muney A. Understanding differences in health behaviors by education. J Health Econ. 2010;29(1):1–28. Epub 2009/12/08. doi: 10.1016/j.jhealeco.2009.10.003.

23. Drageset S, Lindstrøm TC. Coping with a possible breast cancer diagnosis: demographic factors and social support. Journal of Advanced Nursing. 2005;51(3):217–26. doi: doi.org/10.1111/j.1365-2648.2005.03495.x.

24. De-Nour AK. Prediction of adjustment to chronic hemodialysis. Psychonephrology. 1981;1:117–31.

25. Katapodi MC, Facione NC, Miaskowski C, Dodd MJ, Waters C. The influence of social support on breast cancer screening in a multicultural community sample. Oncol Nurs Forum. 2002;29(5):845–52. Epub 2002/06/12. doi: 10.1188/02.Onf.845-852.

26. Chant R, Zoellner BP. Teaching content methods in a high school PDS: navigating curricular tensions. Northwest Journal of Teacher Education. 2023;18(1). doi: doi.org/10.15760/nwjte.2023.18.1.3

27. Khalid M, Akhter. M, Hashmi A. Teaching styles of secondary school English teachers and learning styles of their students and relationship of teaching learning style match with students’ achievement. Bulletin of Education and Research. 2017;39(3):203–20.

28. Zhao X, Kadono M, Kranzler EC, Pavisic I, Miles S, Maher M, et al. Message fatigue and COVID-19 vaccine booster uptake in the United States. Journal of Health Communication. 2024;29(1):61–71. doi: 10.1080/10810730.2023.2282036.

29. Fletcher R, Nielsen RK. Are people incidentally exposed to news on social media? A comparative analysis. New Media & Society. 2018;20(7):2450–68. doi: 10.1177/1461444817724170.

30. Commodari E, La Rosa VL, Coniglio MA. Health risk perceptions in the era of the new coronavirus: are the Italian people ready for a novel virus? A cross-sectional study on perceived personal and comparative susceptibility for infectious diseases. Public Health. 2020;187:8–14. doi: doi.org/10.1016/j.puhe.2020.07.036.

31. Duncan LA, Schaller M. Prejudicial attitudes toward older adults may be exaggerated when people feel vulnerable to infectious disease: Evidence and implications. Analyses of Social Issues and Public Policy. 2009;9(1):97–115. doi: doi.org/10.1111/j.1530-2415.2009.01188.x.

32. Mikels JA, Shuster MM, Thai ST, Smith-Ray R, Waugh CE, Roth K, et al. Messages that matter: Age differences in affective responses to framed health messages. Psychology and Aging. 2016;31(4):409–14. doi: 10.1037/pag0000040.

33. Shamaskin AM, Mikels JA, Reed AE. Getting the message across: Age differences in the positive and negative framing of health care messages. Psychology and Aging. 2010;25(3):746–51. doi: 10.1037/a0018431.

34. Pereira D, Fillol J, Moura P. Young people learning from digital media outside of school: the informal meets the formal. Comunicar: Media Education Research Journal. 2019;27(58):41–50.

35. Boulianne S, Theocharis Y. Young people, digital media, and engagement: A meta-analysis of research. Social science computer review. 2020;38(2):111–27.

36. Livingstone S, Bober M, Helsper EJ. Active participation or just more information? Young people’s take-up of opportunities to act and interact on the Internet. Information, Community & Society. 2005;8(3):287–314. doi: doi.org/10.1080/13691180500259103.

37. Willoughby JF, L’Engle KL. Influence of perceived interactivity of a sexual health text message service on young people’s attitudes, satisfaction and repeat use. Health education research. 2015;30(6):996–1003. doi: 10.1093/her/cyv056.

38. Guillory J, Henes A, Farrelly MC, Fiacco L, Alam I, Curry L, et al. Awareness of and receptivity to the fresh empire tobacco public education campaign among hip hop youth. Journal of Adolescent Health. 2020;66(3):301–7. doi: 10.1016/j.jadohealth.2019.09.005.

39. Guo M, Ganz O, Cruse B, Navarro M, Wagner D, Tate B, et al. Keeping it fresh with hip-hop teens: Promising targeting strategies for delivering public health messages to hard-to-reach audiences. Health Promotion Practice. 2020;21(1_suppl):61S–71S.

40. More KR, More C, Burd KA, Mentzou A, Phillips LA. Does messaging matter? A registered report on appearance-versus health-based message framing in exercise appeals targeted towards women. Psychology of Sport and Exercise. 2024;70:102555. doi: doi.org/10.1016/j.psychsport.2023.102555.

41. So J, Alam N. Predictors and effects of anti-obesity message fatigue: A thought-listing analysis. Health Communication. 2019;34(7):755–63. doi: 10.1080/10410236.2018.1434736.

42. Chen PH, Huang SM, Lai JC, Lin WL. Determinants of health-seeking behavior toward Chinese or Western medicine in Taiwan: An analysis of biobank research database. Complement Ther Clin Pract. 2022;48:101592. Epub 2022/04/20. doi: 10.1016/j.ctcp.2022.101592.

43. Kuan YC, Yen DJ, Yiu CH, Lin YY, Kwan SY, Chen C, et al. Treatment-seeking behavior of people with epilepsy in Taiwan: A preliminary study. Epilepsy Behav. 2011;22(2):308–12. Epub 2011/08/05. doi: 10.1016/j.yebeh.2011.06.034.

44. McKenzie SK, Collings S, Jenkin G, River J. Masculinity, social connectedness, and mental health: Men’s diverse patterns of practice. Am J Mens Health. 2018;12(5):1247–61. Epub 2018/05/01. doi: 10.1177/1557988318772732.

45. Gough B, Novikova I. Mental health, men and culture: how do sociocultural constructions of masculinities relate to men’s mental health help-seeking behaviour in the WHO European Region? Copenhagen: World Health Organization. Regional Office for Europe; 2020.

46. Weinstein ND. Unrealistic optimism about susceptibility to health problems: conclusions from a community-wide sample. J Behav Med. 1987;10(5):481–500. Epub 1987/10/01. doi: 10.1007/bf00846146.

47. Prentice KJ, Gold JM, Carpenter WT, Jr. Optimistic bias in the perception of personal risk: patterns in schizophrenia. Am J Psychiatry. 2005;162(3):507–12. Epub 2005/03/03. doi: 10.1176/appi.ajp.162.3.507.

48. Özaltun E. Overconfidence, self-knowledge, and self-improvement. Palgrave Communications. 2017;3(1):42. doi: 10.1057/s41599-017-0049-5.

